# Time-to-retraction and likelihood of evidence contamination (VITALITY Extension I): a retrospective cohort analysis

**DOI:** 10.64898/2026.02.20.26346631

**Authors:** Yuan Tian, Zhen Peng, Suhail A. Doi, Luis Furuya-Kanamori, Hong Cao, Lifeng Lin, Haitao Chu, Yoon Loke, Ben W Mol, Su Golder, Sunita Vohra, Chang Xu

## Abstract

**Background:** The number of problematic randomized clinical trials (RCTs) has risen sharply in recent decades, posing serious challenges to the integrity of the healthcare evidence ecosystem.

**Objective:** To investigate whether retraction of problematic RCTs could reduce evidence contamination.

**Design:** Retrospective cohort study

**Setting:** A secondary analysis of the VITALITY Study database.

**Participants:** 1,330 retracted RCTs with 847 systematic reviews.

**Measurements:** The difference in the median number (and its interquartile, IQR) of contamination before and after retraction. The association between time-to-retraction and likelihood of evidence contamination.

**Results:** Among these retracted RCTs, 426 led to evidence contamination, resulting in 1,106 contamination events (251 after retraction vs. 855 before retraction). The time interval between RCT publication and first contamination ranged from 0.2 to 30.9 years, with a median of 3.3 years (95% CI: 3.0 to 3.9). The median number of contaminated systematic reviews was lower after retraction than before retraction (0, IQR: 0 to 1 vs. 1, IQR: 1 to 2, *P* < 0.01). Compared with trials retracted more than 7.5 years after publication, those retracted between 1.0 and 1.8 years (OR = 0.70, 95% CI: 0.60 to 0.80) and retracted within 1.0 year (OR = 0.69, 95% CI: 0.60 to 0.80) were associated with lower likelihood of evidence contamination.

**Limitations:** Only assessed contaminated systematic reviews with quantitative synthesis and limited to retracted RCTs.

**Conclusions:** Retracting problematic RCTs can significantly reduce evidence contamination, and faster retraction was associated with less contamination. To safeguard the integrity of the evidence ecosystem, academic journals should act promptly in the retraction of problematic studies to minimize their downstream impact.

**Primary Funding Sources:** The National Natural Science Foundation of China (72204003, 72574229)

## Introduction

In clinical research, randomized controlled trials (RCTs) are considered to be the most rigorous study design for establishing causal relationships between health interventions and outcomes [1, 2]. The strict randomization process utilized in RCTs is intended to ensure that the comparator groups are “exchangeable”, meaning that each group possesses the same baseline risk for specific conditions [3]. This methodological design allows researchers to minimize potential biases, such as confounding [4]. Due to this distinctive feature, RCTs have gained widespread acceptance for pre-marketing evaluation of new investigational drugs, medical devices, and health-related interventions [5-7].

Despite their methodological strengths, RCTs are not immune to bias. Sub-optimal design, conduct, and analysis can introduce biases such as selection bias and measurement bias, resulting in deviations of estimated effects from the true values [8, 9]. Over the past fifty years, researchers in clinical epidemiology have made significant efforts to address these biases [9-11]. Various statistical and epidemiological methods are available to measure and deal with the issue of bias [12-15]. However, all these efforts rely on one fundamental assumption: that RCTs are conducted with integrity, and without fabrication or falsification by competent researchers [16, 17].

In recent decades, an increasing number of retracted RCTs have been documented due to the questionable nature of their data and practices, raising significant concerns and contributing to a crisis of trust within the scientific community [18-20]. Research misconduct has emerged as a critical threat in the field of clinical epidemiology [20, 21]. The negative consequences of retracted RCTs extend beyond the evidence they present; their substantial dissemination rates significantly amplify their influence, for example, causes evidence contamination on evidence ecosystem [22, 23].

How to mitigate the impact of potentially problematic studies remains a pressing research question for clinical epidemiology [24-26]. Research has documented that the number of citations diminished rapidly after the retraction of problematic studies [27-29]. Moreover, retracted studies received an annual average of 2.5 citations, lower than that of 3.6 citations for non-retracted studies [30]. These findings demonstrated a fact that timely retraction of problematic studies would mitigate their citation. While evidence contamination differs conceptually from citation, it remains unclear whether retraction could mitigate its impact on evidence ecosystem.

To answer this question, we conducted a retrospective study to bridge this gap, based on the VITALITY Study cohort, to compare the number of evidence contamination in systematic reviews for problematic trials before and after their retraction.

## Methods

### Study Design

This study employed a retrospective cohort design, and was reported according to the STROBE checklist for cohort studies [31]. We utilized the VITALITY Study I database, which encompassed a cohort of retracted trials obtained from the Retraction Watch database, and a subsequent cohort of systematic reviews that quantitatively synthesized these retracted trials identified through forward citation searching [23]. Since evidence from these retracted trials was synthesized into the systematic reviews, evidence contamination occurred, and these reviews were therefore identified as contaminated. The cohort of retracted trials and the cohort of contaminated systematic reviews were then used to investigate the association between trial retraction and evidence contamination.

### Database

The VITALITY Study I database, established on 21 November 2024, comprises 1,330 retracted trials and 847 systematic reviews that quantitatively synthesized retracted trials. In brief, retracted RCTs from the Retraction Watch database (http://retractiondatabase.org/) up to November 5, 2024 were screened and collected. We then performed forward citation searching for each retracted trial via Google Scholar and Scopus to identify systematic reviews that quantitatively synthesized these trials. We excluded records that provided limited information (e.g., conference abstracts), replicated other meta-analyses (e.g., umbrella reviews), were published in non-English languages, or had been retracted. Details of the search strategy and screening methodology have been reported elsewhere [23].

### Data collection

For the retracted trials, the following data were directly extracted from the Retraction Watch database: Digital Object Identifier (DOI), title, journal of publication, journal quartiles, date of publication, date of retraction, retraction initiator (e.g., author), authors of the retraction. We also collected information from the main texts of retracted trials, including trial registration status (e.g., yes, no), data sharing statement (e.g., yes, no), study center (e.g., single) and funding sources (e.g., industry supported).

For systematic reviews contaminated by these trials, the following data were collected: DOI, title, date of publication, date of submission, date of the literature search, number of retracted trials synthesized, number of trials included in each meta-analysis, and metadata. Date of publication, submission and literature search were collected from the review reports or journal websites by one author (Y.T) and cross-verified by another (Z.P). All other data were obtained directly from VITALITY Study dataset.

### Exposure and Outcomes

The primary outcome was the difference in the median number of contaminated systematic reviews before and after retraction across trials. The secondary outcome was the likelihood of evidence contamination, with the exposure of interest defined as the time interval between trial publication and retraction (i.e., time-to-retraction). We also estimated the time-to-first contamination, which was defined as the time interval between trial publication and the first publication of the systematic review contaminated by the trial. The number of contaminated systematic reviews was counted at the trial level, as a single systematic review could be contaminated by multiple trials. For example, if both Trial A and Trial B contaminated the same systematic review, this was counted as one contamination event for Trial A and one for Trial B.

### Statistical analysis

The baseline characteristics of the eligible trials were summarized using proportions and median values with interquartile ranges (IQR), according to the nature of the data. The median time-to-first contamination with its quartiles was estimated via Kaplan-Meier analysis [32]. The differences in the number of contamination events after versus before retraction were assessed using the Mann–Whitney U test [33].

The determination of whether the contamination occurred before or after the retraction was based on two time points, namely, the retraction date of problematic trials (T0) as well as the start date of systematic reviews (T1). We used the start date (T1) instead of the publication date (T2) of systematic reviews to account for the publication delay (Tm), during which new retractions unknown to the authors may occur. We estimated Tm as the median time from submission to publication and estimated T1 as *T*2 −*Tm*. Contamination was classified as before retraction if T1 < T0 and after retraction otherwise. When the submission time was unavailable, the literature search date was used as a proxy.

A generalized linear model was employed to examine the association between time-to-retraction and the likelihood of evidence contamination, with cluster-robust standard errors to account for potential clustering of retractions by the same authors. We stratified time-to-retraction into four categories in terms of its 25%, 50% and 75% percentiles. To identify potential confounders, a directed acyclic graph (DAG) was employed to depict causal relationships between time-to-retraction (exposure) and evidence contamination (outcome) (Supplementary **Figure S1**). Based on the DAG, the following covariates were identified as important confounders and were adjusted: journal ranking (Q1, Q2, Q3, Q4, and no information), retraction initiators (editor initiated vs. author initiated), source of funding (industry supported, non-profit, no funding, and not reported), and registration status (Yes vs. No).

Sensitivity analyses were conducted by using the 25th and 75th percentiles of the time from submission to publication as alternative publication delays (Tm). Data analysis was undertaken using Stata/SE 18.0 (StataCorp, College Station, TX, USA), with an alpha of 0.05 as the statistical significance level.

## Results

Table 1 presents the baseline characteristics of the 1,330 retracted trials. Most of the trials were retracted either after 2022 (43.7%, n = 581) or between 2011 and 2022 (51.9%, n = 691). The majority of retractions were initiated by the journals (85.0%, n = 1,131), while only 9.2% (n = 123) were initiated by trial authors. Amongst the 1,330 retracted trials, 426 (32.0%) led to evidence contamination, resulting in 1,106 contamination events (251 after retraction vs. 855 before retraction), and embodied in a total of 847 systematic reviews. Among the 847 contaminated systematic reviews, the majority of reviews (89.0%, n = 754) were contaminated by a single retracted trial, while 11.0% (n = 93) were contaminated by at least two retracted trials.

**Table 1.** Basic characteristics of 1,330 retracted trials.

|  | Summary of retracted trials |  |
| --- | --- | --- |
|  | Trials cause evidence contamination (n=426) | Trials did not cause evidence contamination (n=904) |
| **Characteristics** |  |  |
| **Time-to-retraction, Median (IQR)** | 6.03 (1.90 to 13.28) | 1.35 (0.93 to 3.72) |
| **Year of retraction, Number (%)** |  |  |
| 2010 and before | 20 (4.7%) | 38 (4.2%) |
| 2011 to 2022 | 294 (69.0%) | 397 (43.9%) |
| 2023 and after | 112 (26.3%) | 469 (51.9%) |
| **Trial registration, Number (%)** |  |  |
| Yes | 109 (25.6%) | 140 (15.5%) |
| No | 317 (74.4%) | 764 (84.5%) |
| **Data-related retractions, Number (%)** |  |  |
| Yes | 300 (70.4%) | 642 (71.0%) |
| No | 126 (29.6%) | 262 (29.0%) |
| **Funding source, Number (%)** |  |  |
| Industry supported | 10 (2.4%) | 24 (2.7%) |
| Non-industry supported | 88 (20.7%) | 224 (24.8%) |
| Not funded | 42 (9.9%) | 64 (7.1%) |
| Not reported | 286 (67.1%) | 592 (65.5%) |
| **Data sharing statement, Number (%)** |  |  |
| Yes | 54 (12.7%) | 443 (49.0%) |
| No | 372 (87.3%) | 461 (51.0%) |
| **Retraction procedure, Number (%)** |  |  |
| Author initiated | 46 (10.8%) | 77 (8.5%) |
| Journal initiated | 359 (84.3%) | 772 (85.4%) |
| Author and journal initiated | 9 (2.1%) | 17 (1.9%) |
| Unclear | 12 (2.8%) | 38 (4.2%) |
| **Center information, Number (%)** |  |  |
| Single center | 172 (40.4%) | 312 (34.5%) |
| Multiple centers | 67 (15.7%) | 79 (8.7%) |
| Missing | 187 (43.9%) | 513 (56.8%) |
| **Journal quartiles, Number (%)** |  |  |
| Q1 | 188 (44.1%) | 228 (25.2%) |
| Q2 | 138 (32.4%) | 258 (28.5%) |
| Q3 | 51 (12.0%) | 107 (11.8%) |
| Q4 | 11 (2.6%) | 30 (3.3%) |
| No information | 38 (8.9%) | 281 (31.1%) |

For the 847 systematic reviews, the median time from submission to publication for was 0.5 year (IQR: 0.3 to 0.8), which was used to address the publication delays. The time-to-first contamination ranged from 0.2 to 30.9 years, with a median of 3.3 years (95% CI: 3.0 to 3.9). The 10th percentile of time-to-first contamination was 0.9 year (95% CI: 0.8 to 1.2), 25th percentile was 1.7 years (95% CI: 1.5 to 1.9), and 75th percentile was 6.6 years (95% CI: 6.1 to 7.4), see Figure 1.

**Figure 1.**
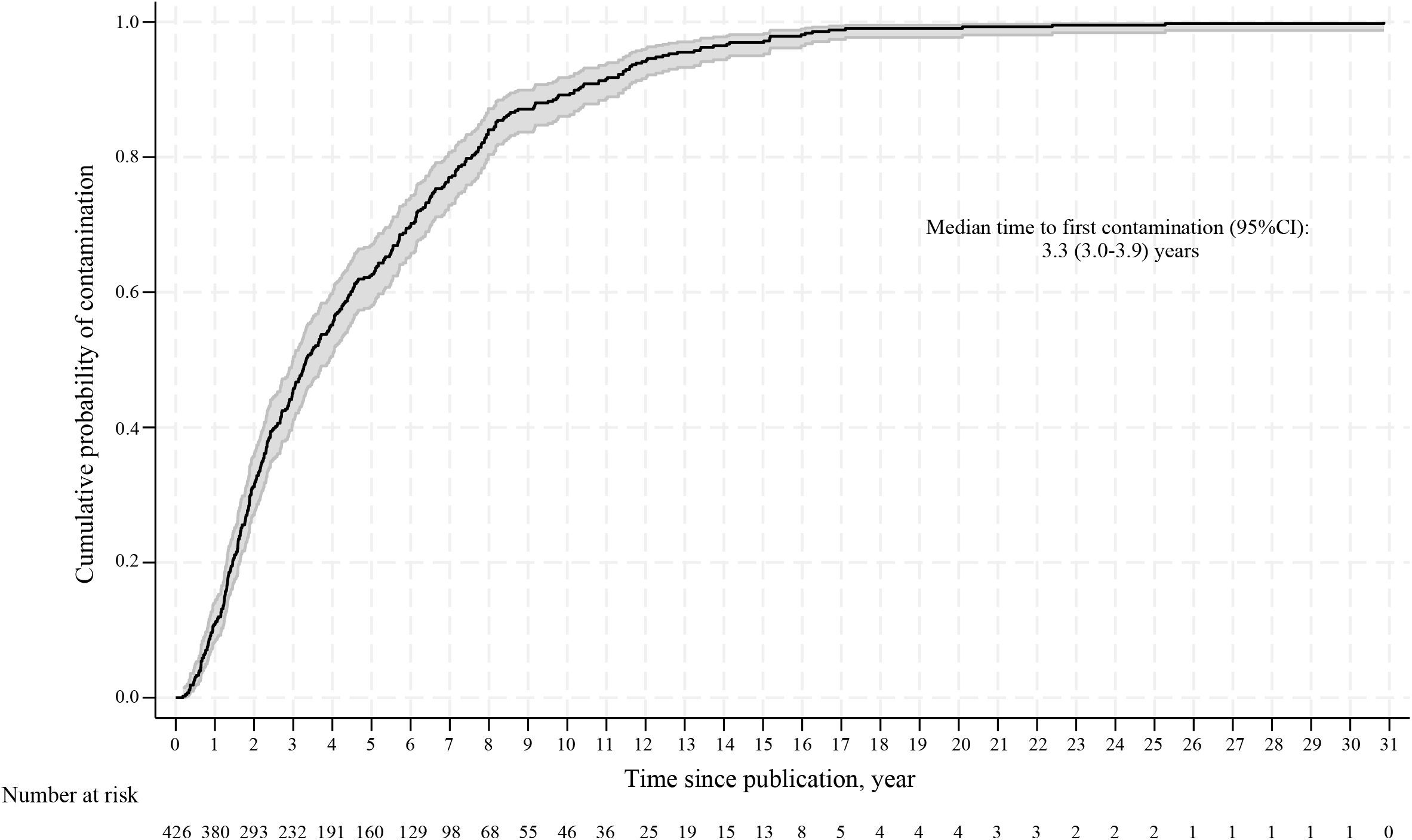
Analysis of time to first contamination for systematic reviews (n = 426).

**Figure 2.**
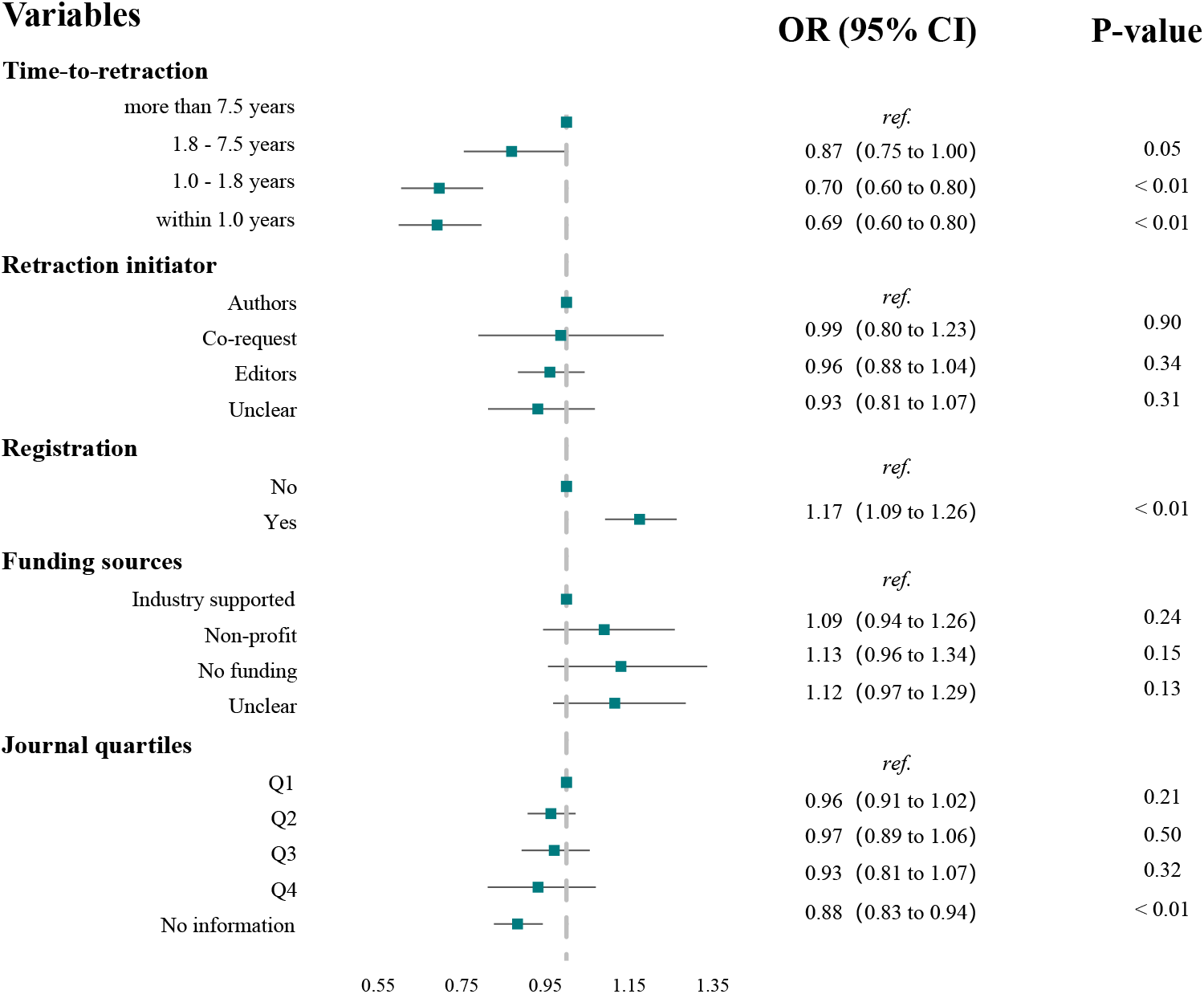
Association between features of retracted trials and evidence contamination

### Events of contamination after and before retraction

The median number of contaminated systematic reviews was 0 (IQR: 0 to 1) after retraction as compared to before retraction 1 (IQR: 1 to 2) (*P* < 0.01). Sensitivity analyses were performed by using the 25th and 75th percentiles of the time from submission to publication as alternative publication delays (Tm). Under the 25th percentile, median number of contaminated systematic reviews was 0 (IQR: 0 to 1) after retraction compared to 1 (IQR: 1 to 2) before retraction (*P* < 0.01). Under the 75th percentile, the median number of contaminated systematic reviews was 0 (IQR: 0 to 1) after retraction compared to 2 (IQR: 1 to 3) before retraction (*P* < 0.01).

### Association between time-to-retraction and evidence contamination

For the 1,330 retracted trials, 426 retracted trials led to evidence contamination. Compared with trials retracted more than 7.5 years after publication, those retracted between 1 and 1.8 years (OR = 0.70, 95% CI: 0.60 to 0.80, p < 0.01) and retracted within 1.0 year (OR = 0.69, 95% CI: 0.60 to 0.80) were associated with significant lower likelihood of evidence contamination. However, those retracted between 1.8 and 7.5 years (OR = 0.87, 95%CI: 0.75 to 1.00, p = 0.05) were marginally associated with lower likelihood of evidence contamination.

### Post-hoc analyses

We noticed that among the 426 retracted trials, 95 (22.3%) resulted in contamination only after their retraction. Therefore, post-hoc analyses were conducted to explore the potential mechanisms.

These 95 trials contaminated 124 systematic reviews (14.6%) involving 460 meta-analyses. The time-to-first contamination ranged from 0.5 to 23.9 years, with a median of 1.8 years (IQR: 1.3 to 2.5) (see Supplementary **Figure S2**). The median publication-to-retraction time for these trials was 1.1 years (IQR: 0.6 to 2.8), dramatically shorter than that median of the whole sample (5.9 years, IQR: 1.9 to 13.1). Among the 441 meta-analyses that could be replicated, exclusion of retracted trials resulted in a change in the significance of *P* value in 15.0% (95%CI: 11.8% to 18.6%; n = 66), which is comparable to the whole sample (16.0%, 95%CI: 14.2% to 17.9%).

## Discussion

This study investigated the association between trial retraction and the extent of evidence contamination in systematic reviews. We found that 90% of the evidence contamination occurred after 0.9 years following publication of the retracted trials, 75% after 1.7 years, and 50% after 3.3 years. Whilst problematic trials continued to cause evidence contamination after their retraction, retraction of such problematic trials can significantly reduce evidence contamination, and a faster retraction was associated with less contamination. These findings yield two key implications for the scientific community: 1) while retracting problematic trials is a necessary step to mitigate their harms, it is not sufficient as a standalone solution; 2) comprehensive strategies are needed to minimize their negative impacts, for example, instituting widespread warnings of their risks, and developing or updating guidelines for conducting and reporting systematic reviews to identify and remove retracted trials.

Although the retraction of problematic trials is essential for maintaining the trustworthiness of the evidence ecosystem, prior research indicates that the median time from the initiation of an investigation by journals to issue an outcome (e.g., retraction) can extend to 38 months [23, 34]. Such delays may reflect the complexity of misconduct investigations, inconsistencies in journal retraction policies, and limitations of post-publication assessment mechanisms [29]. However, empirical evidence on how delays in retraction affect evidence contamination remains limited. In this study, we addressed this knowledge gap and found that retracting problematic trials within one year of publication could reduce contamination by 90%. This finding underscores the need for journals to accelerate early detection and retraction processes to mitigate the propagation of contaminated evidence.

It is notable that we observed about 22% of the problematic trials caused evidence contamination only after their retraction. Based on our post hoc analysis, these trials were retracted more rapidly than the whole sample (1.1 years vs. 5.9 years), and 0.5 years before the first contamination occurred. This suggested that authors of the systematic reviews contaminated by these trials probably have already known the retraction status while continue to included them. We further investigated whether potential conflicts of interest exist among related meta-analyses but failed to observe a signal—the impact of these retracted trials on the contaminated meta-analyses was comparable to the whole sample (15.0% vs. 16.0% in terms of change on the significance). Therefore, a more likely possibility would be owing to the missing labeling of the retraction status of such trials. Previous research has documented that less than half of the retracted studies were electronically marked as retracted [35]. In this case, review authors remained unaware of the status and therefore included them into the evidence synthesis.

To our knowledge, this is the first study to investigate the impact of time-to-retraction on evidence contamination in systematic reviews. We used nearly all retracted trials that were retrieved from Retraction Watch (up to November 2024) in the analysis, as an effort to maximize the representativeness. In addition, we tracked the citations of these retracted trials via both the Google Scholar and Scopus databases to capture systematic reviews contaminated by these retracted trials, which ensured our data covered the majority of these reviews. The process of citation screening and data extraction were also rigorously conducted, with two rounds of training before the formal analysis began. In the main analysis, we employed a before-and-after design that allowed us to control for all potential confounding factors, thereby minimizing potential bias. All these efforts strengthen the reliability of the findings.

However, there are several limitations that should be highlighted. First, our current study did not account for the contamination of retracted trials in systematic reviews without meta-analysis, systematic reviews of multiple-arm comparisons, umbrella reviews, scoping reviews, meta-analyses of proportions, etc., which are expected to underestimate the risk of evidence contamination. In addition, this study did not take retracted non-randomized studies of intervention or observational studies into consideration, and thus the findings may not reflect contamination arising from these study types. Both would substantially underestimate the extent of evidence contamination and therefore bias our results. Further studies should address such issues using more comprehensive dataset.

In conclusion, retracting a problematic trial can reduce evidence contamination, and faster retraction was associated with less contamination of systematic reviews by problematic trials. Academic journals should act promptly on problematic studies to prevent negative impacts on the evidence ecosystem. Additional comprehensive strategies beyond timely retraction are also needed to minimize the ripple effects of retracted trials.

## Data Availability

Statistical code and data set: the original data can be found at https://osf.io/gb5ed.

## Acknowledgment

None.

## Financial Support

This study was supported by the National Natural Science Foundation of China (72204003, 72574229), the institutional funding by Shanghai Eastern Hepatobiliary Surgery Hospital of Navy Medical University (‘TengFei Project’, TF2024YZRH03), and Hefei Comprehensive National Science Center (0301035204), and ShangHai Shenkang Hospital Development Center (SHDC12025626). Suhail Doi was supported by Program Grant #NPRP-BSRA01-0406-210030 from the Qatar National Research Fund.

## Disclosure

Disclosure forms are available with the article online

## Reproducible Research Statement

Study protocol: Not available. Statistical code and data set: the original data can be found at https://osf.io/gb5ed.

## Authors’ contributions

Conception and design: CX; Manuscript drafting: CX, TY, ZP; Data collection: YT, ZP; Data analysis and result interpretation: TY, ZP, CX; Statistical guidance: LL, CH, CX; Methodology guidance: SD, BM, LFK, CX; Manuscript editing: CX, ZHF, YT, JHF, ZP, YL, SD, LFK, LL, CH, BM, SG, SV. All authors have read and approved the manuscript.

## Notes

### Competing Interest Statement

The authors have declared no competing interest.

### Summary of Updates

Figure 1 and Figure 2 upload with the more clear files. Supplemental files upload.

