## Supplementary Figure S1 for "Time-to-retraction and likelihood of evidence contamination (VITALITY Extension I): a retrospective cohort analysis"

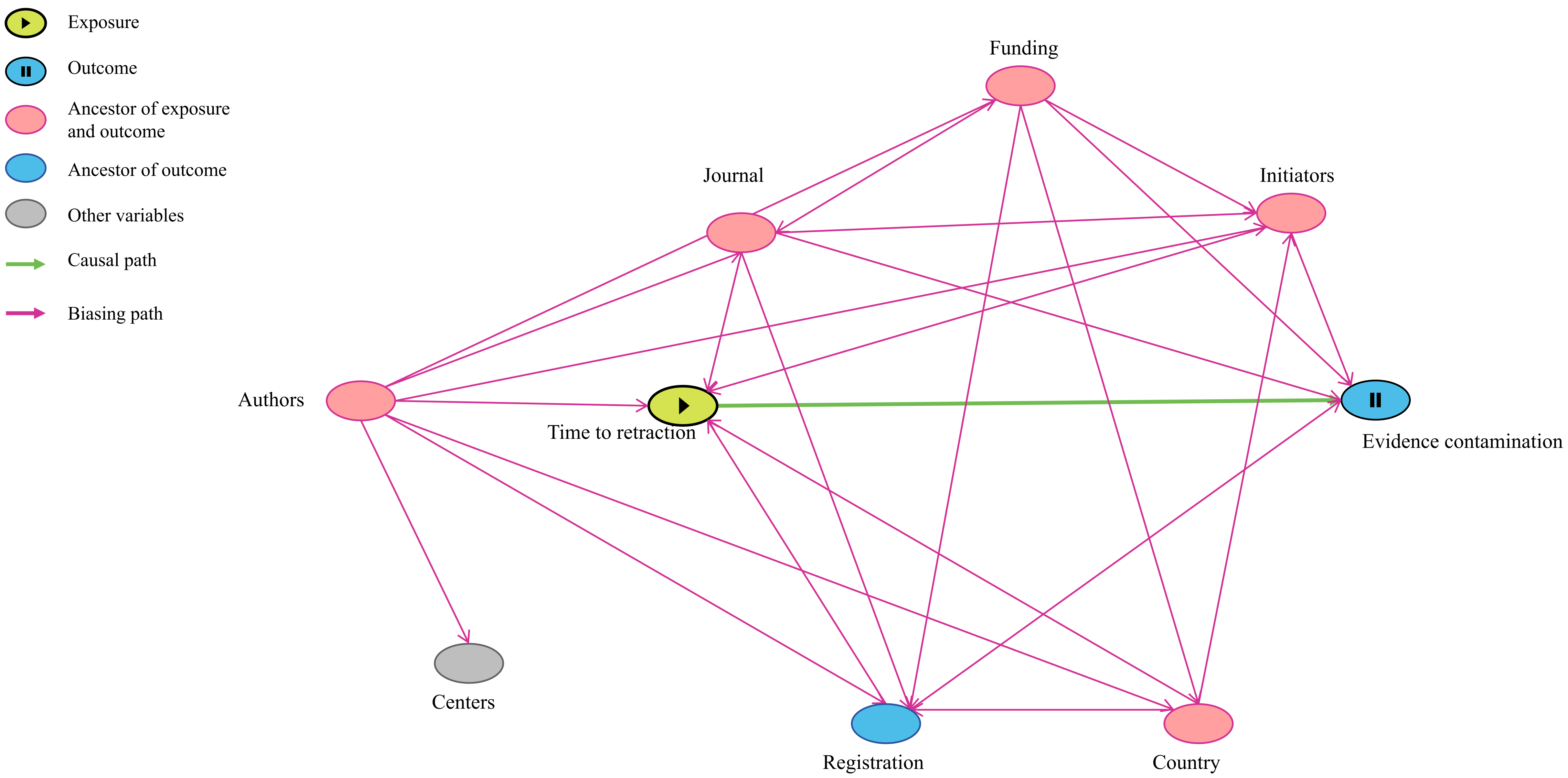


Minimal sufficient adjustment sets for estimating the total/direct effect of Time on Contamination: (1) *Funding, registration, initiators, journal*; (2) Author, Country, Funding, initiators, journal;
