## Supplementary figures and images for "Time-to-retraction and likelihood of evidence contamination (VITALITY Extension I): a retrospective cohort analysis"

### Supplementary Figure S2

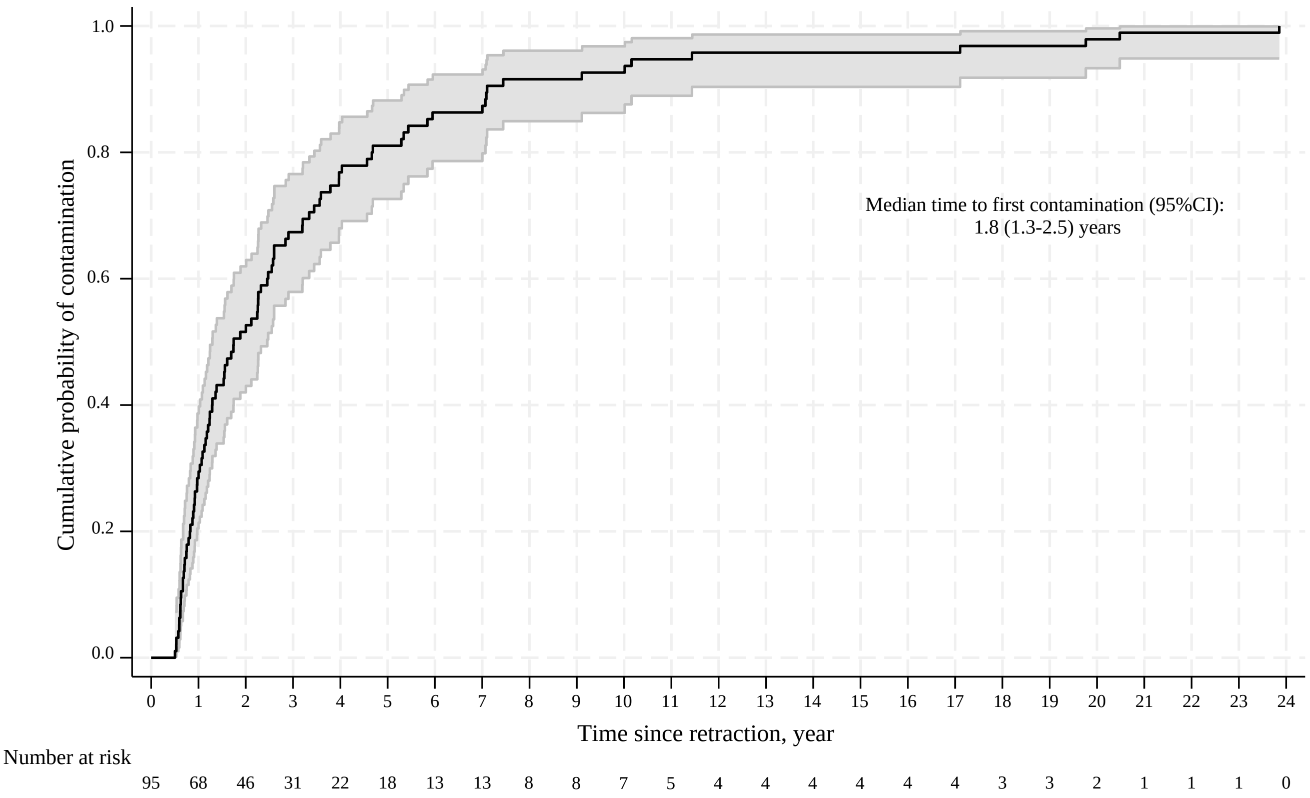

### Supplementary Figure S3

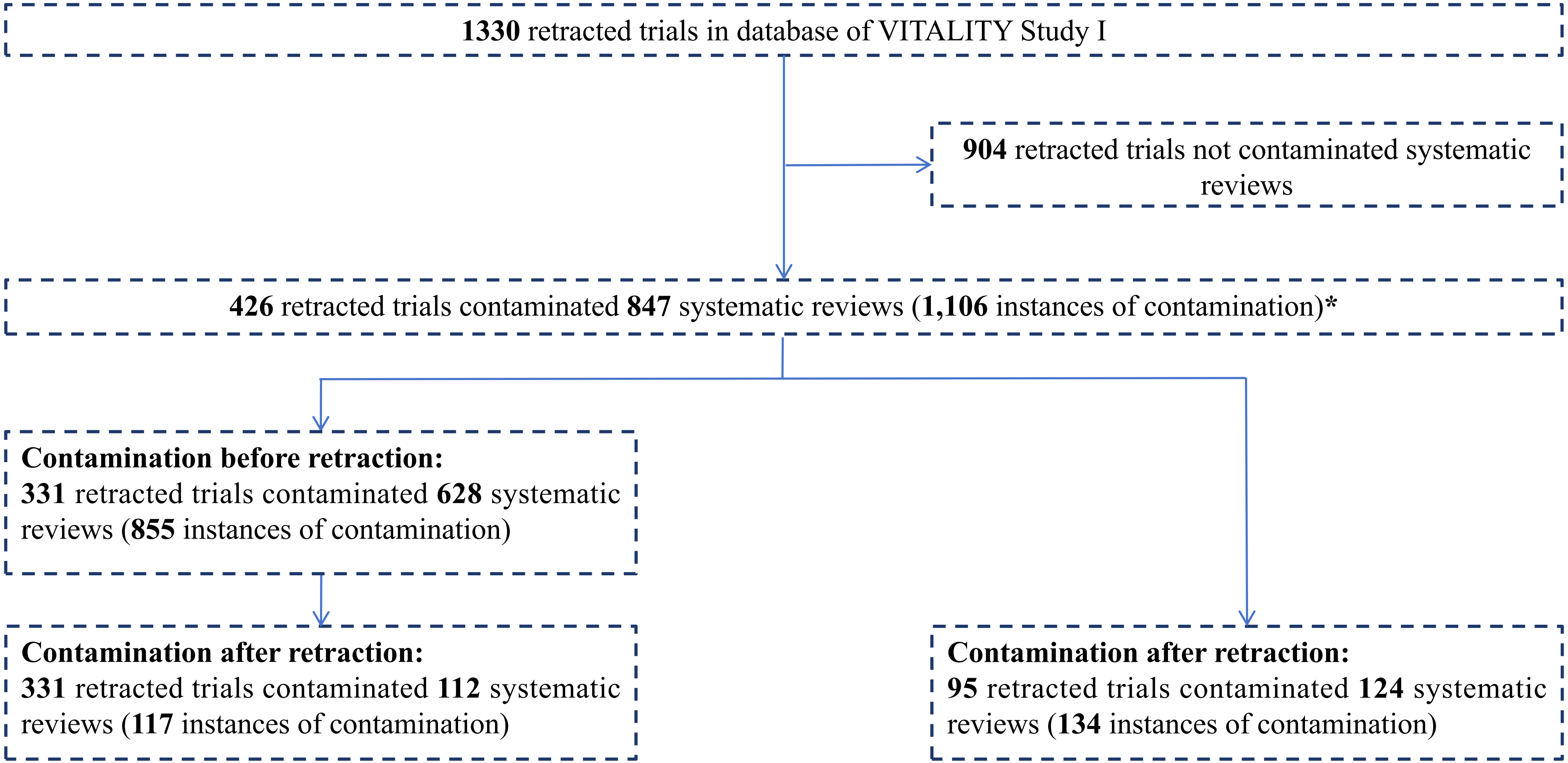


***Contamination was tracked until 5 November 2024**
