## Supplementary file for "Time-to-retraction and likelihood of evidence contamination (VITALITY Extension I): a retrospective cohort analysis"

**STROBE Statement**—Checklist of items that should be included in reports of ***cohort studies***

|  | Item No | Recommendation | Page No |
| --- | --- | --- | --- |
| **Title and abstract** | 1 | (*a*) Indicate the study’s design with a commonly used term in the title or the abstract | Page 1 |
|  |  | (*b*) Provide in the abstract an informative and balanced summary of what was done and what was found | Page 2 |
| Introduction | | | |
| Background/rationale | 2 | Explain the scientific background and rationale for the investigation being reported | Page 3 |
| Objectives | 3 | State specific objectives, including any prespecified hypotheses | Page 4 |
| Methods | | | |
| Study design | 4 | Present key elements of study design early in the paper | Page 4 |
| Setting | 5 | Describe the setting, locations, and relevant dates, including periods of recruitment, exposure, follow-up, and data collection | Page 4 |
| Participants | 6 | (*a*) Give the eligibility criteria, and the sources and methods of selection of participants. Describe methods of follow-up | Page 4 |
|  |  | (*b*) For matched studies, give matching criteria and number of exposed and unexposed | Not applicable |
| Variables | 7 | Clearly define all outcomes, exposures, predictors, potential confounders, and effect modifiers. Give diagnostic criteria, if applicable | Page 5 |
| Data sources/ measurement | 8* | For each variable of interest, give sources of data and details of methods of assessment (measurement). Describe comparability of assessment methods if there is more than one group | Page 5 |
| Bias | 9 | Describe any efforts to address potential sources of bias | Page 6 |
| Study size | 10 | Explain how the study size was arrived at | Page 4 |
| Quantitative variables | 11 | Explain how quantitative variables were handled in the analyses. If applicable, describe which groupings were chosen and why | Page 6 |
| Statistical methods | 12 | (*a*) Describe all statistical methods, including those used to control for confounding | Page 6 |
|  |  | (*b*) Describe any methods used to examine subgroups and interactions | Not applicable |
|  |  | (*c*) Explain how missing data were addressed | Page 6 |
|  |  | (*d*) If applicable, explain how loss to follow-up was addressed | Not applicable |
|  |  | (*e*) Describe any sensitivity analyses | Page 6 |
| Results | |  | |
| Participants | 13* | (a) Report numbers of individuals at each stage of study—eg numbers potentially eligible, examined for eligibility, confirmed eligible, included in the study, completing follow-up, and analysed | Page 7 |
|  |  | (b) Give reasons for non-participation at each stage | Supplementary Figure S3 |
|  |  | (c) Consider use of a flow diagram | Supplementary Figure S3 |
| Descriptive data | 14* | (a) Give characteristics of study participants (eg demographic, clinical, social) and information on exposures and potential confounders | Page 16 |
|  |  | (b) Indicate number of participants with missing data for each variable of interest | Page 6 |
|  |  | (c) Summarise follow-up time (eg, average and total amount) | Not applicable |
| Outcome data | 15* | Report numbers of outcome events or summary measures over time | Page 7 |

| Main results | 16 | (*a*) Give unadjusted estimates and, if applicable, confounder-adjusted estimates and their precision (eg, 95% confidence interval). Make clear which confounders were adjusted for and why they were included | Page 6 |
| --- | --- | --- | --- |
|  |  | (*b*) Report category boundaries when continuous variables were categorized | Page 6 |
|  |  | (*c*) If relevant, consider translating estimates of relative risk into absolute risk for a meaningful time period | Not applicable |
| Other analyses | 17 | Report other analyses done—eg analyses of subgroups and interactions, and sensitivity analyses | Page 6 |
| Discussion | | | |
| Key results | 18 | Summarise key results with reference to study objectives | Page 8 |
| Limitations | 19 | Discuss limitations of the study, taking into account sources of potential bias or imprecision. Discuss both direction and magnitude of any potential bias | Page 10 |
| Interpretation | 20 | Give a cautious overall interpretation of results considering objectives, limitations, multiplicity of analyses, results from similar studies, and other relevant evidence | Page 8-10 |
| Generalisability | 21 | Discuss the generalisability (external validity) of the study results | Page 10 |
| Other information | | | |
| Funding | 22 | Give the source of funding and the role of the funders for the present study and, if applicable, for the original study on which the present article is based | Page 11 |

*Give information separately for exposed and unexposed groups.

Note: An Explanation and Elaboration article discusses each checklist item and gives methodological background and published examples of transparent reporting. The STROBE checklist is best used in conjunction with this article (freely available on the Web sites of PLoS Medicine at http://www.plosmedicine.org/, Annals of Internal Medicine at http://www.annals.org/, and Epidemiology at http://www.epidem.com/). Information on the STROBE Initiative is available at http://www.strobe-statement.org.

**DAG plot for association between time to retraction and evidence contamination**

We used DAG plot to identify potential confounders that needed to be addressed. To obtain causal paths between exposure (time-to-retraction) and outcome (evidence contamination), we reviewed previous studies and taken these variables into consideration: (1) journals quartiles (Q1, Q2, Q3, Q4 and No information); (2) country; (3) funding (Industry funded, Non-profit, No funding and Not reported); (4) initiators of retraction (Editor, Author, Co-request and Unclear); (5) registration (Yes, No); (6) centers (Single, Multiply and Unclear); and (7) cluster of author.

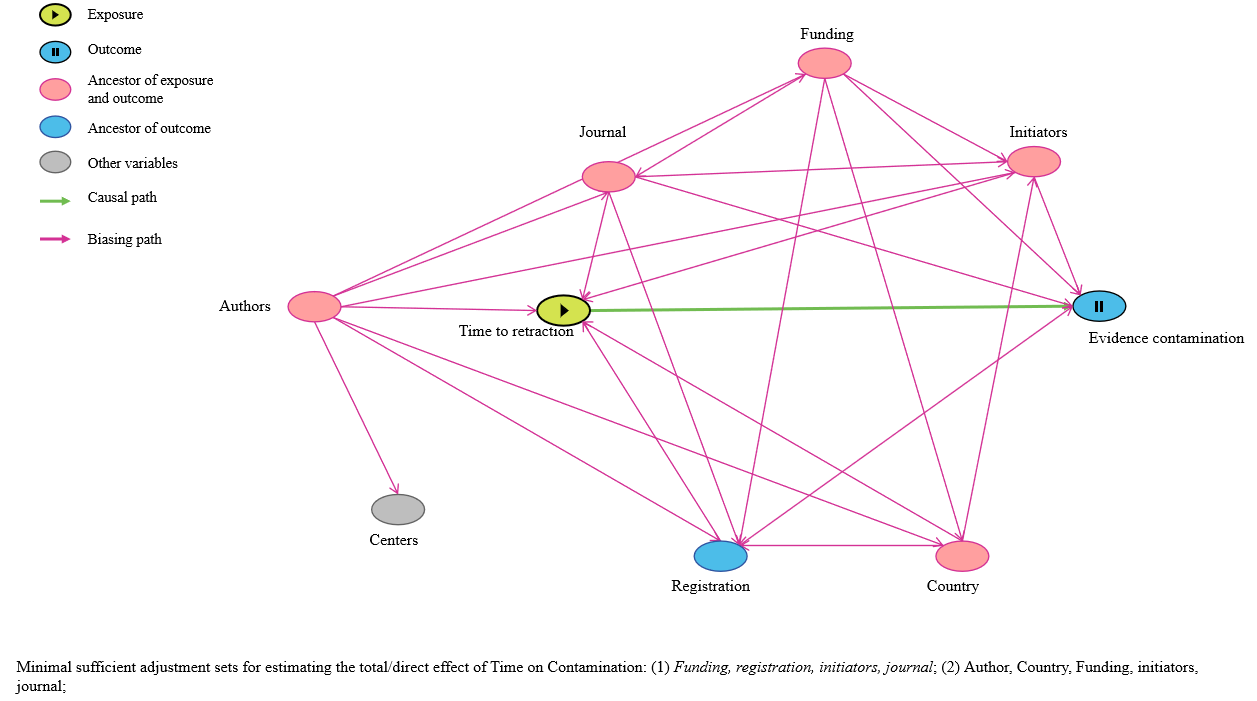
**Figure S1.** The DAG plot for identifying potential confounders for regression analysis

**The DAG plot code** (http://dagitty.net/).

dag {

bb="0,0,1,1"

author [pos="0.112,0.455"]

center [pos="0.127,0.596"]

country [pos="0.648,0.705"]

evidence [outcome,pos="0.733,0.395"]

funding [pos="0.509,0.152"]

initiators [pos="0.681,0.235"]

journal [pos="0.323,0.212"]

reg [pos="0.428,0.717"]

time [exposure,pos="0.293,0.462"]

author -> center

author -> country

author -> funding

author -> initiators

author -> journal

author -> reg

author -> time

country -> initiators

country -> reg

country -> time

funding -> country

funding -> evidence

funding -> initiators

funding -> journal

funding -> reg

initiators -> evidence

initiators -> time

journal -> evidence

journal -> initiators

journal -> reg

journal -> time

reg -> evidence

time -> evidence

}

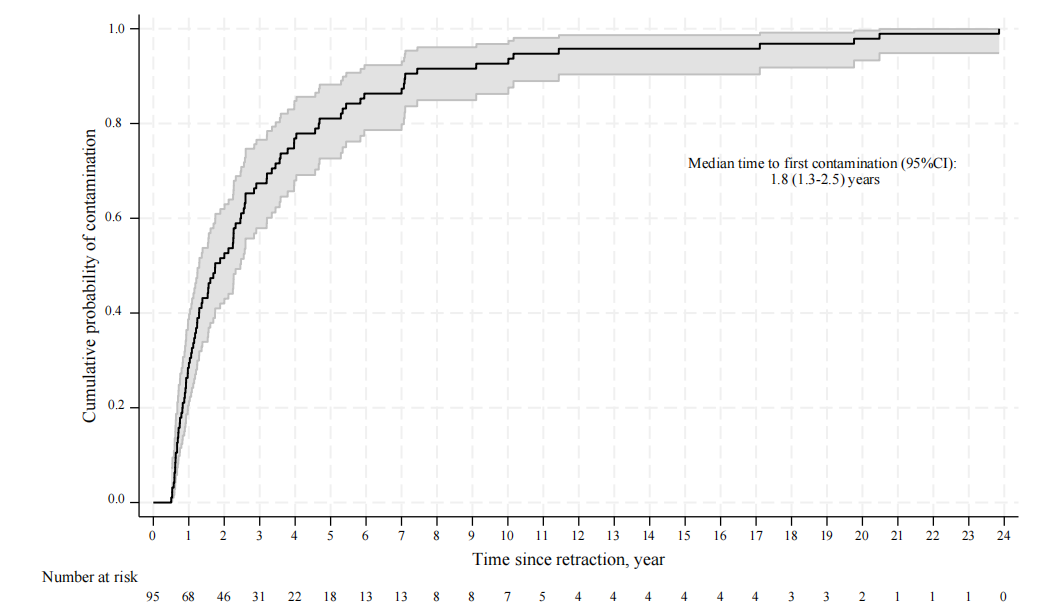
**Figure S2.** Time to first contamination for retracted RCTs resulted in contamination only after their retraction.

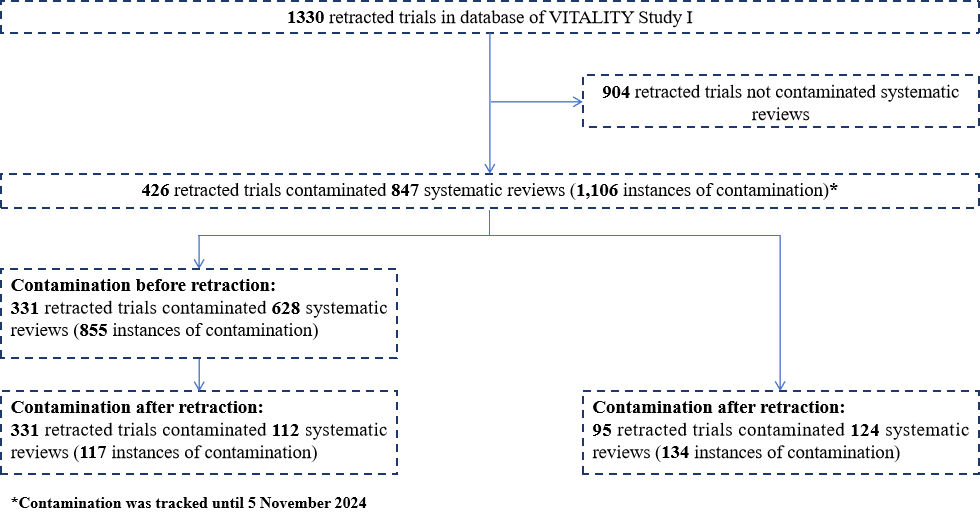
**Figure S3**. Flowchart for contaminated systematic reviews.

**Statistical code for Stata**

******baseline information******

gen reg = .

replace reg = 1 if registration == "Y"

replace reg = 0 if registration == "N"

table reg

gen fund = .

replace fund = 2 if funding == "No"

replace fund = 3 if funding == "Not reported"

replace fund = 1 if funding == "Institution and Government" | funding == "Government" | funding == "Institution" | funding == "Others" | funding == "others"

replace fund = 0 if funding == "Industry" | funding == "Industry and Government" | funding == "Industry and Institution"

table fund

gen initiator = .

replace initiator = 0 if retractioninitiator == "Author"

replace initiator = 1 if retractioninitiator == "Co-request"

replace initiator = 2 if retractioninitiator == "Editor"

replace initiator = 3 if retractioninitiator == "Unclear"

gen jour = .

replace jour = 1 if journalquartiles == "Q1"

replace jour = 2 if journalquartiles == "Q2"

replace jour = 3 if journalquartiles == "Q3"

replace jour = 4 if journalquartiles == "Q4"

replace jour = 5 if journalquartiles == "NA"

summarize survival, detail

******Regression analysis for contamination and spline curved******

glm contam i.survival i.initiator i.reg i.fund i.jour,vce (cluster indicator) eform

******time to first contamination******

stset timeoffirstcontaminationyear

stci

stci, p(10)

stci, p(25)

stci, p(75)
